# Dynamical Behavior Analysis of 2-control Strategies on Tuberculosis Model

**DOI:** 10.64898/2026.01.13.26343993

**Authors:** M. A. Salek, J. Nayeem, Muhammad Sajjad Hossain, M. Humayun Kabir

**Affiliations:** Mathematical and Computation Biology (MCB) Group, Department of Mathematics, Jahangirnagar University, Dhaka, Bangladesh; Department of Arts and Sciences, Ahsanullah University of Science and Technology, Bangladesh

**Keywords:** Tuberculosis, Optimal control, Pontryagain’s maximum principle, reproduction number, sensitivity analysis

## Abstract

To characterize tuberculosis transmission and assess the impact of important interventions, a data-driven SEITR TB model is created. The potential for disease persistence has been calculated using the basic reproduction number. To determine the factors most significantly affecting the spread of tuberculosis, stability and sensitivity analyses are conducted. Strengthened treatment measures and optimized distancing significantly lower infection levels, according to numerical simulations. The Least Squares Fitting technique is used to validate real epidemiological data with a model solution. And the results emphasize that the best combinations of social distancing and treatment not only reduce the number of infections but also provide a cost-effective strategy for public health planning. Additionally, two numerical techniques, namely Pearson correlation and Partial Rank Correlation Coefficients (PRCC), are utilized to assess the sensitivity of model parameters. It is noted that the outcomes of these two methods are in agreeable comparison with one another regarding sensitivity analysis.

## 1 Introduction

Tuberculosis (TB), caused by the bacterium *Mycobacterium tuberculosis (MTB)*, is a major global health issue that mainly affects the lungs [**?**]. While lung infections are most common, the bacteria can spread to other organs through the bloodstream, leading to extrapulmonary TB, which is usually non-infectious [**?**]. Active TB shows symptoms like a longlasting cough, chest pain, fever, weight loss, and night sweats. It spreads through airborne droplets. Doctors typically diagnose TB with skin tests a few weeks after exposure, even though symptoms may not show up until later. Researchers employ an SEITR compartmental model to comprehend the transmission of tuberculosis. This aids in the formulation of effective and efficient public health strategies [**?**, **?**]. Understanding how stable and sensitive a person is helps decide where to put treatment and social distancing efforts. This can help communities control TB better in the long run [**?**, **?**]. A lot of the current TB models aren’t very accurate because they don’t take into account things like seasonal changes, reinfection, or the best ways to treat the disease [**?**, **?**].

Optimal control theory has been shown to be a useful tool for finding ways to reduce disease while making the best use of available resources [**?**]. This method is especially helpful for keeping TB under control in places where there aren’t many health care resources [**?**]. By systematically simulating disease progression under various conditions, we can pinpoint critical transmission drivers and assess the sensitivity of interventions [**?**]. Mathematical modeling has also been useful for figuring out how effective and cost-effective TB control programs will be in the long run [**?**]. Additionally, behavior factors and longer treatment needs have been included to improve realism and lower the chances of disease reactivation [**?**].

In this study, we used a SEITR model by introducing clear exposed and treated compartments to better capture the progression of TB and its control dynamics [**?**, **?**]. We integrated optimal control strategies to evaluate the performance of interventions in different epidemiological scenarios and to reduce the overall disease burden [**?**]. Previous research has shown that TB elimination is possible when the basic reproduction number is below one; otherwise, persistence is expected [**?**, **?**]. Further studies have highlighted the need to combine vaccination, treatment, and preventive measures to meet biomedical and economic goals [**?**, **?**]. This proposed model adds to the existing literature by offering a structured mathematical basis for assessing effective and sustainable TB control strategies [**?**].The following are the key components of this study:

a. In what ways can stability analysis identify the threshold conditions that naturally suppress the spread of tuberculosis within a population?
b. When treatment and distancing techniques are adjusted, which epidemiological parameters have the most impact on the dynamics of tuberculosis?
c. How much can the prioritization of scarce public health resources in tuberculosis control programs be influenced by sensitivity analysis?
d. How can the predictive power of SEITR-based tuberculosis models be improved by explicitly include exposed and treated compartments?
e. Is it possible for mathematically optimized control systems to achieve sustainable tuberculosis reduction more effectively than traditional intervention approaches?

This work demonstrates how these issues are resolved by a mathematically rigorous SEITR model that includes stability, sensitivity, and optimal control evaluations. The results are presented to facilitate evidence-based decision-making for effective and sustainable tuberculosis prevention.

The article is structured as follows: Section 2 provides a comprehensive description of the baseline tuberculosis model, emphasizing its mathematical properties and dynamics like [5] in Section 3. Section 4 presents the outcomes of numerical simulations related to the baseline model and the optimal control problem. And Section 5 summarizes the key conclusions and suggestions of the study.

## 2 Governing Model

The SEITR model is a compartmental approach that shows susceptible (*S*) people who are healthy but at risk of getting TB, exposed (*E*) people who have been exposed to TB but are not yet infectious, infectious (*I*) people who have active TB and can spread the disease to others, treated (*T* ) people who are getting treatment for TB, and recovered (*R*) people, as shown in Figure 1. We integrate control variables into the SEITR model to accommodate actual measures designed to reduce the transmission of tuberculosis (TB). Optimal control theory allows us to identify the most effective strategies for minimizing both the number of infectious individuals and the costs associated with these interventions. Here, we define two key control variables such as Distance Control (*u*_1_(*t*)) and Treatment Control (*u*_2_(*t*)) in this model as[**?**]. The non-linear differential equations, which reflect the dynamics between compartments of ordinary differential equations as in [**?**], are given below. The description of the model parameters is provided in Table 1 of the Appendix.

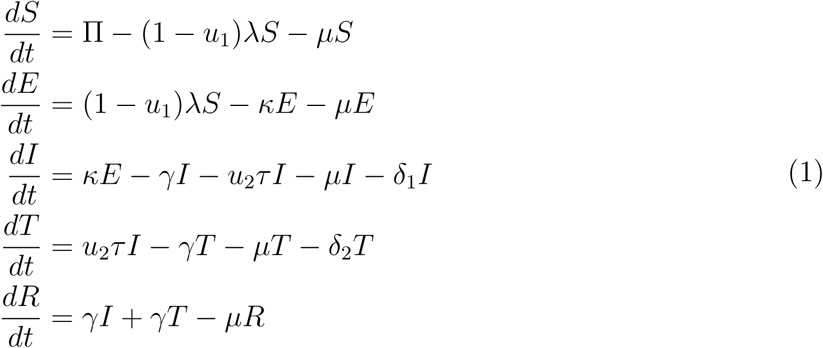

where,

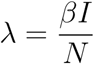

and the non-negative initial conditions are, *S*_0_ ≥ 0*, E*_0_ ≥ 0*, I*_0_ ≥ 0*, T*_0_ ≥ 0*, R*_0_ ≥ 0.

**Figure 1:**
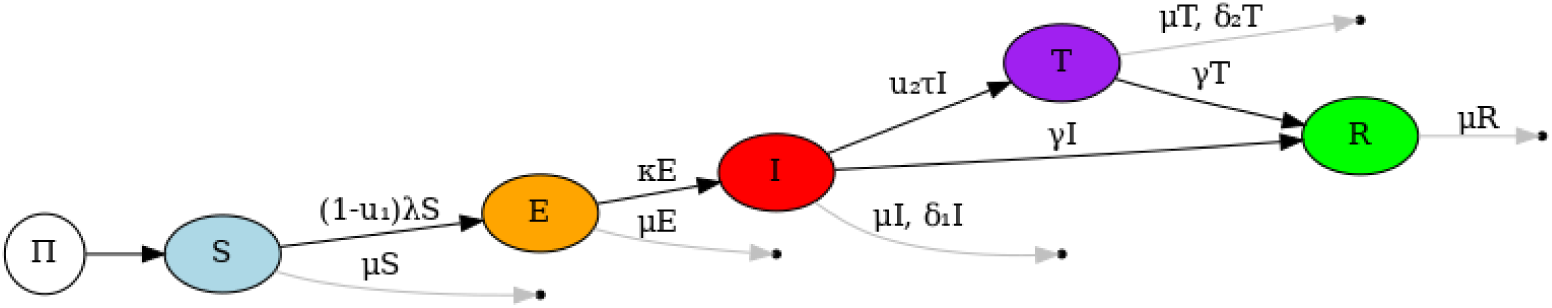
Diagram scheme of TB transmission with control strategies.

**Table 1:** An overview of the model’s variable components.

| Parameters | Biological Meaning | Values |
| --- | --- | --- |
| $\mu$ | Natural death rate | 0.02 [?] |
| $\eta$ | Acute infection rate | 1.2 (Estimated) |
| $\delta_1$ | Diseases induced death rate of acute infected class | 0.1 (Estimated) |
| $\delta_2$ | Diseases induced death rate of treatment class | 0.1 (Estimated) |
| $\kappa$ | Progression rate of exposed class | 0.85 [?] |
| $\tau$ | Treatment rate | 0.8 [?] |
| $\gamma$ | Recovery rate | 0.23 [?] |
| $\Pi$ | Constant recruitment rate | 2000 (Estimated) |
| $\beta$ | Transmission rate | 0.45 [?] |

Also *u*_1_(*t*) is the distancing control rate and the coefficient 1−*u*_1_(*t*) represents the prevention efforts to protect people in susceptible compartment from contacting infected individuals. And *u*_2_(*t*) represents the enhancement treatment effort for individuals in the infectious compartment.

## 3 Materials and Methods

### 3.1 Disease Free Equilibrium (DFE) of the model

The model (**??**) exhibits a DFE, which is established by equating the right-hand sides of its equations to zero [**?**].

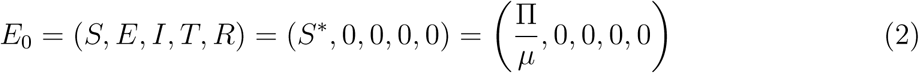

The stability of the DFE, *E*_0_ will be analyzed by next-generation method [**?**]. Presented below are the non-negative matrix *P* , which denotes the new infection terms, and *V* is the non-singular *M* -matrix, which relates to the remaining transfer terms.

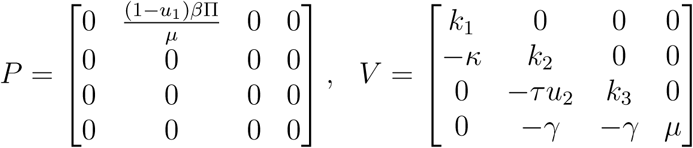

*R*_0_ = *ρ*(*PV* ^−1^), where *ρ* signifies the spectral radius (the largest eigenvalue in absolute value) of the next generation matrix *PV* ^−1^. Consequently, it can be inferred that

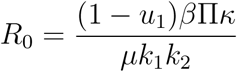

where, *k*_1_ = *κ* + *µ, k*_2_ = *γ* + *u*_2_*τ* + *µ* + *δ*_1_*, k*_3_ = *γ* + *µ* + *δ*_2_.

### 3.2 Positivity and Boundedness of the model

The given model equations are

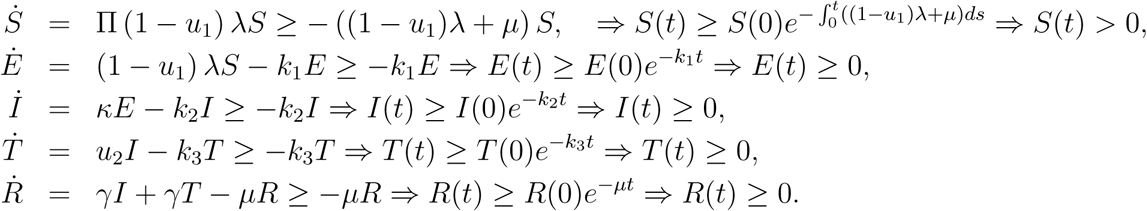

Since, *N* = *S* + *E* + *I* + *T* + *R*, then from (**??**) we have,

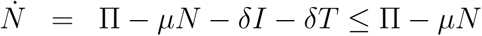

Now,

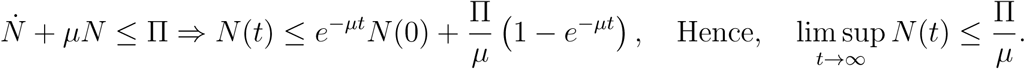

**Theorem 1** *The solution of the system is positively bounded for all* (*S*(0)*, E*(0)*, I*(0)*, T* (0)*, R*(0)) ∈ *R*^5^_+_ *and also define for the positive value of time t*.

### 3.3 Invariant Region

The dynamical transmission of the system (**??**) will be analyzed in the below biological feasible region, 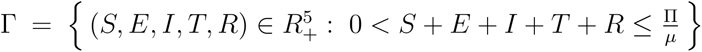. The solution set is uniformly bounded and established the global existence. The solution for our model is continuously dependent on initial data. Therefore, our model is biologically well-proposed.

### 3.4 Objective Function of Optimal Control

The objective function quantifies the trade-off between reducing the infectious population and minimizing intervention costs over a fixed time horizon. It is expressed as:

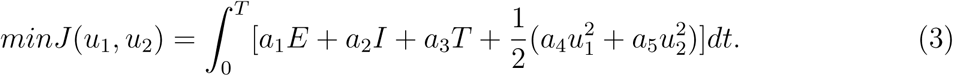

where, *a*_1_*, a*_2_*, a*_3_*, a*_4_ and *a*_5_ are weight parameters and all are positive. The squared terms represent the cost of control measures, assuming they increase quadratically with their intensity.

### 3.5 Characterization of the Optimal Control

By Pontryagin’s Maximum Principle (PMP), which transforms the problem into a system of differential equations involving state variables *S*(*t*)*, E*(*t*)*, I*(*t*)*, T* (*t*) and *R*(*t*) adjoint variables: *λ_S_, λ_E_, λ_I_, λ_T_* and *λ_R_* and Hamiltonian. The Hamiltonian incorporates the objective function and state dynamics as:

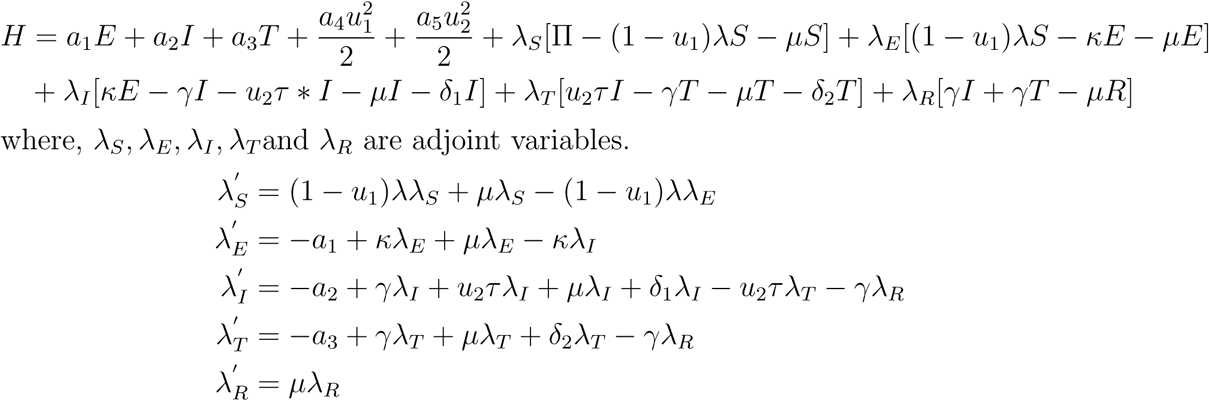

The adjoint variables *λ_i_* satisfying 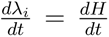 with the transversity conditions, *λ_i_*(*T* ) = 0, *i* = *S, E, I, T, R* .

The optimality condition is given by 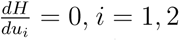. For the control *u*_1_ we have, 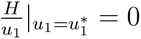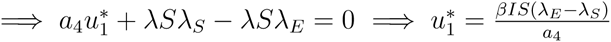

and, for the control *u*_2_ we can write, 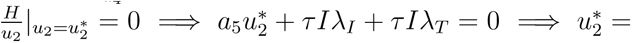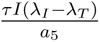

So, we have the controls,

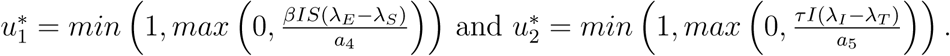

### 3.6 Comparison Study for Infectious and Treatment Population Trajectories using Control Strategies

The long-term effects of control strategies on infected and treated populations over a 25-year period are shown in Figures 2(a) and 2(b). When compared to the uncontrolled situation, a significant decrease in infection levels is seen under controlled conditions, indicating the efficacy of the implemented interventions. By preventing the spread of disease, control techniques significantly reduce the strain on treatment resources, as seen by a corresponding drop in treatment demand.

**Figure 2:**
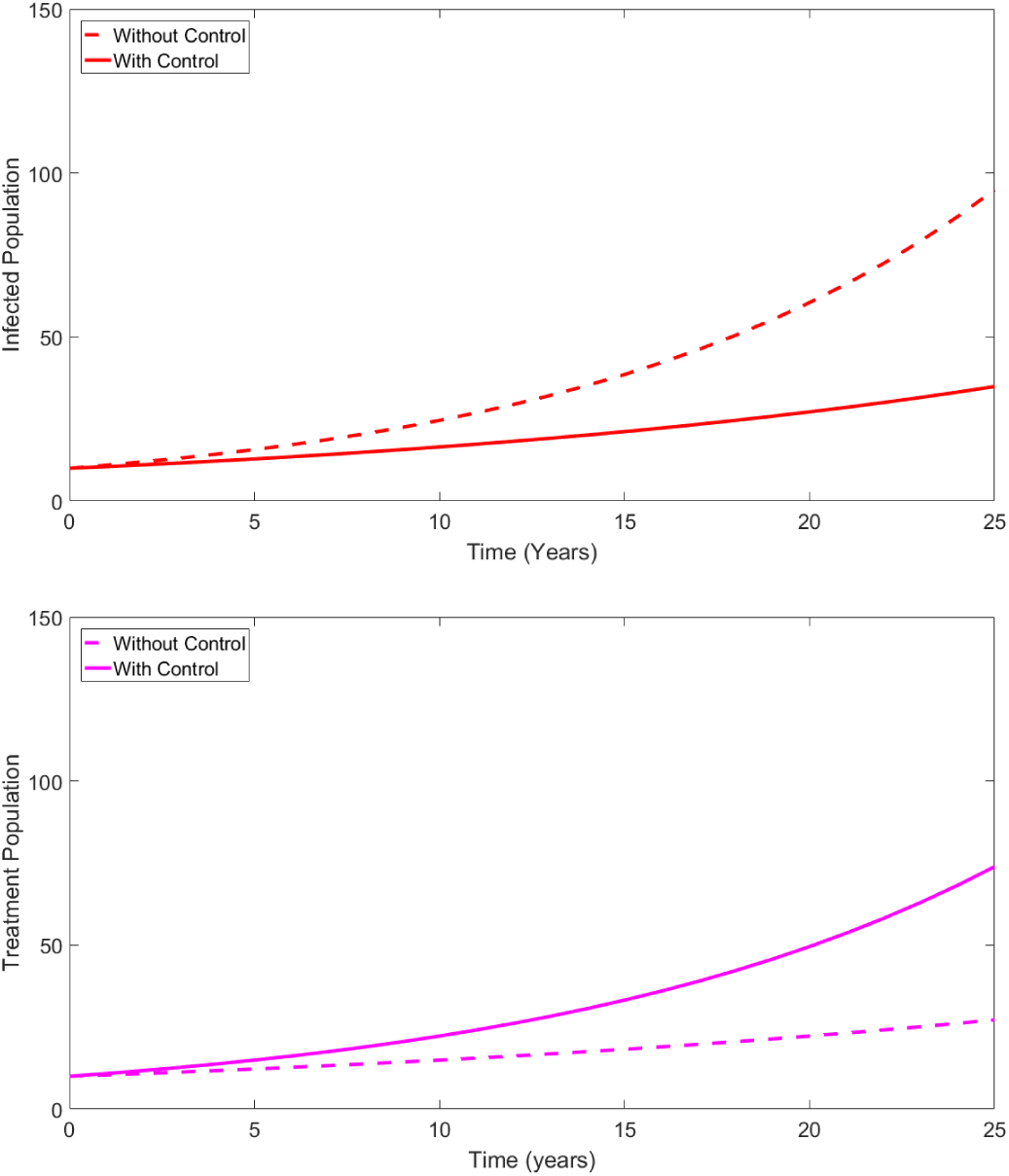
Comparison of infectious (a) and treated population (b) trajectories under controls.

## 4 Numerical Technique

Numerical simulations are performed by discretising the SEITR system and employing typical time-stepping methods to approximate the model states over time. The impact of parameters on model outputs has been assessed by Pearson correlation to measure linear associations between inputs and responses. Furthermore, monotonic effects under concurrent parameter adjustment are analysed using Partial Rank Correlation Coefficients. The consistent outcomes from both methods validated the robustness of the numerical sensitivity analysis and the dependability of parameter ranking.

## 5 Numerical Results and Discussions

### 5.1 Sensitivity Analysis of Different Parameter’s Impact on Infectious Population

The sensitivity analysis presented in the figures highlights the differential impact of varying parameters *β*, *γ*, and *κ* on the infectious population over 50 years. For *β* (Figure 3(a)), which probably stands for the effective contact rate, a +10% increase (dashed red line) causes a big rise in the infectious population compared to the baseline (solid black line). A −10% decrease (dotted blue line) does the opposite, showing that the population is very sensitive to changes in transmission. For *γ* (Figure 3(b)), which shows the recovery rate, a +10% increase (dashed red line) leads to a decrease in the number of people who are sick, since more people recover. On the other hand, a −10% decrease (dotted blue line) leads to a noticeable rise in the number of people who are sick, which shows how important recovery is for stopping the spread.

**Figure 3:**
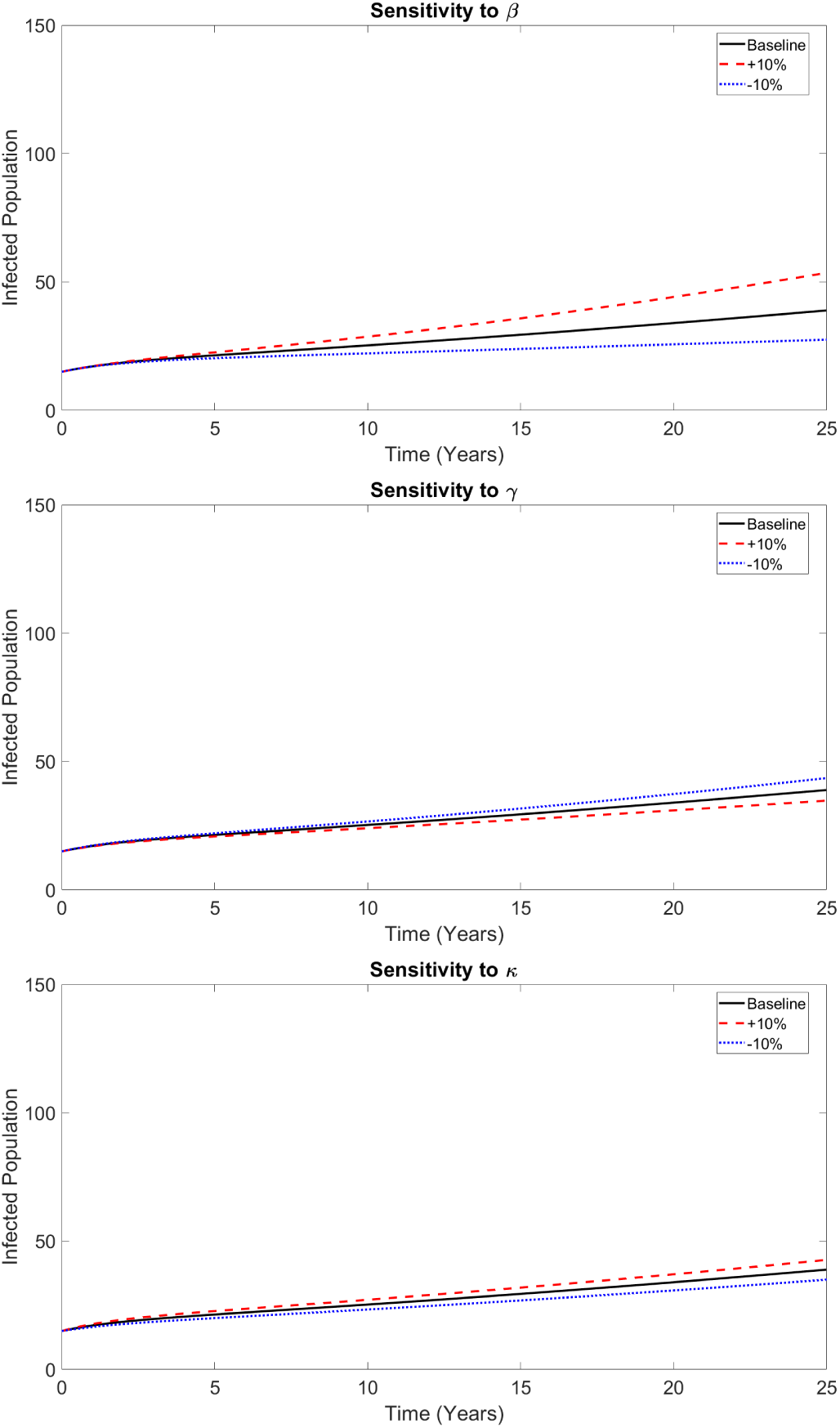
Sensitivity Analysis impact on infectiousness for (a) *β*, (b) *γ*, and (c) *κ*.

Similarly, for *κ* (Figure 3(c)), likely the progression rate from exposed to infectious, a +10% increase (dashed red line) shows an elevated infectious population, and a −10% decrease (dotted blue line) leads to a lower one, though its impact appears less pronounced than that of *β* or *γ*. Overall, these figures demonstrate that the infectious population is most sensitive to changes in the effective contact rate (*β*) and recovery rate (*γ*), emphasizing these parameters as critical targets for intervention strategies aimed at controlling the disease.

### 5.2 Analytic Analysis of Sensitive Parameters

To analyze the sensitivity we defined an explicit formula for *R*_0_ as,

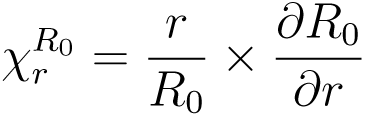

where, *R*_0_ = *R*_0_(*β, κ, γ, τ, δ*_1_*, u*_1_*, u*_2_) and *r* represents a parameter. Some of the key parameters (Mai,2023) of our model (**??**) significantly affect the basic reproduction number *R*_0_ are given in Table 2.

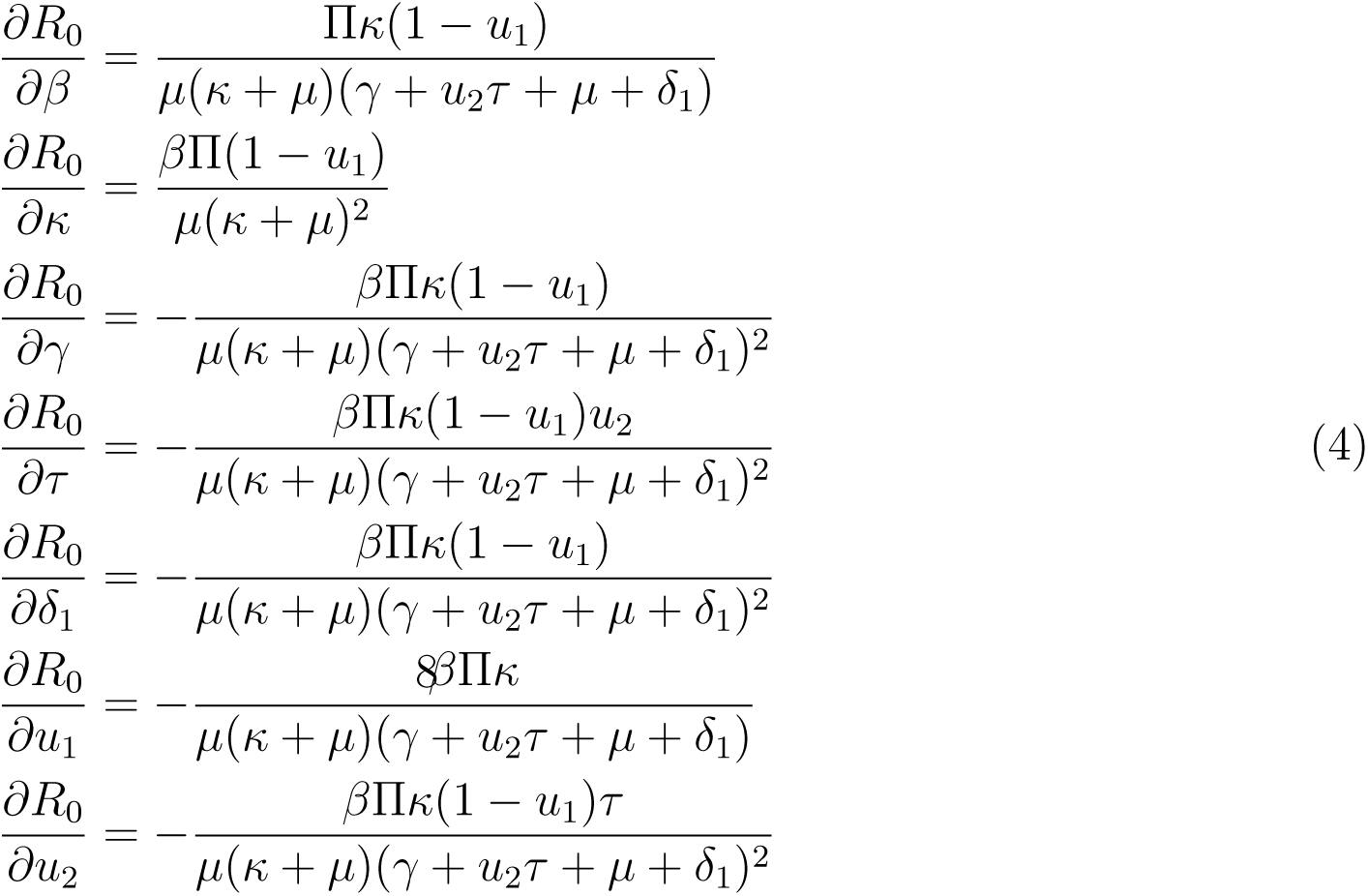

**Table 2:** Indices of sensitivity of *R*_0_ to some model parameters.

| Parameters | Indices of sensitivity |
| --- | --- |
| $\beta$ | 0.5714 |
| $\kappa$ | 0.2605 |
| $\gamma$ | -1.6195 |
| $\tau$ | -0.1728 |
| $\delta_1$ | -0.3457 |
| $u_1$ | -0.5159 |
| $u_2$ | -0.2765 |

Figure 4 analyzed the sensitivity of the basic reproduction number (*R*_0_) to diverse epidemiological and control parameters of the model through scatter plots, regression analysis, Pearson correlation, and Partial Rank Correlation Coefficients (PRCC). The transmission rate (*β*) proved to be the most influential parameter on *R*_0_, demonstrating a slope of 0.49, a Pearson correlation of 0.63, and a PRCC of 0.65, signifying a strong positive correlation. This means that even small changes in the transmission rate can cause a big jump in the reproduction number, which makes it more likely that disease outbreaks will happen. The progression rate from exposed to infected (*κ*) also had a moderate positive effect on *R*_0_, with a slope of 0.22 and a PRCC of 0.21. This shows how important it is to have interventions that slow down the transition to active infection. The treatment control rate (*u*_2_) had the most negative effect on *R*_0_ of all the control-related parameters. Its slope was −0.38, its Pearson correlation was −0.20, and its PRCC was −0.25. This means that improving treatment methods can greatly reduce transmission. The distancing control effort (*u*_1_) and the treatment rate (*τ* ) both had negative slopes of −0.36 and −0.38, respectively. However, their PRCC values were low, at −0.09 and −0.02, respectively. This means that behavioral interventions like distancing can help lower *R*_0_, but their effects might not be very strong on their own if they aren’t used with good case management and treatment. Remarkably, the recovery rate (*γ*) showed an almost insignificant PRCC of (0.04) but a slight positive slope of (0.27), possibly as a result of interaction effects or nonlinear saturation behavior.

**Figure 4:**
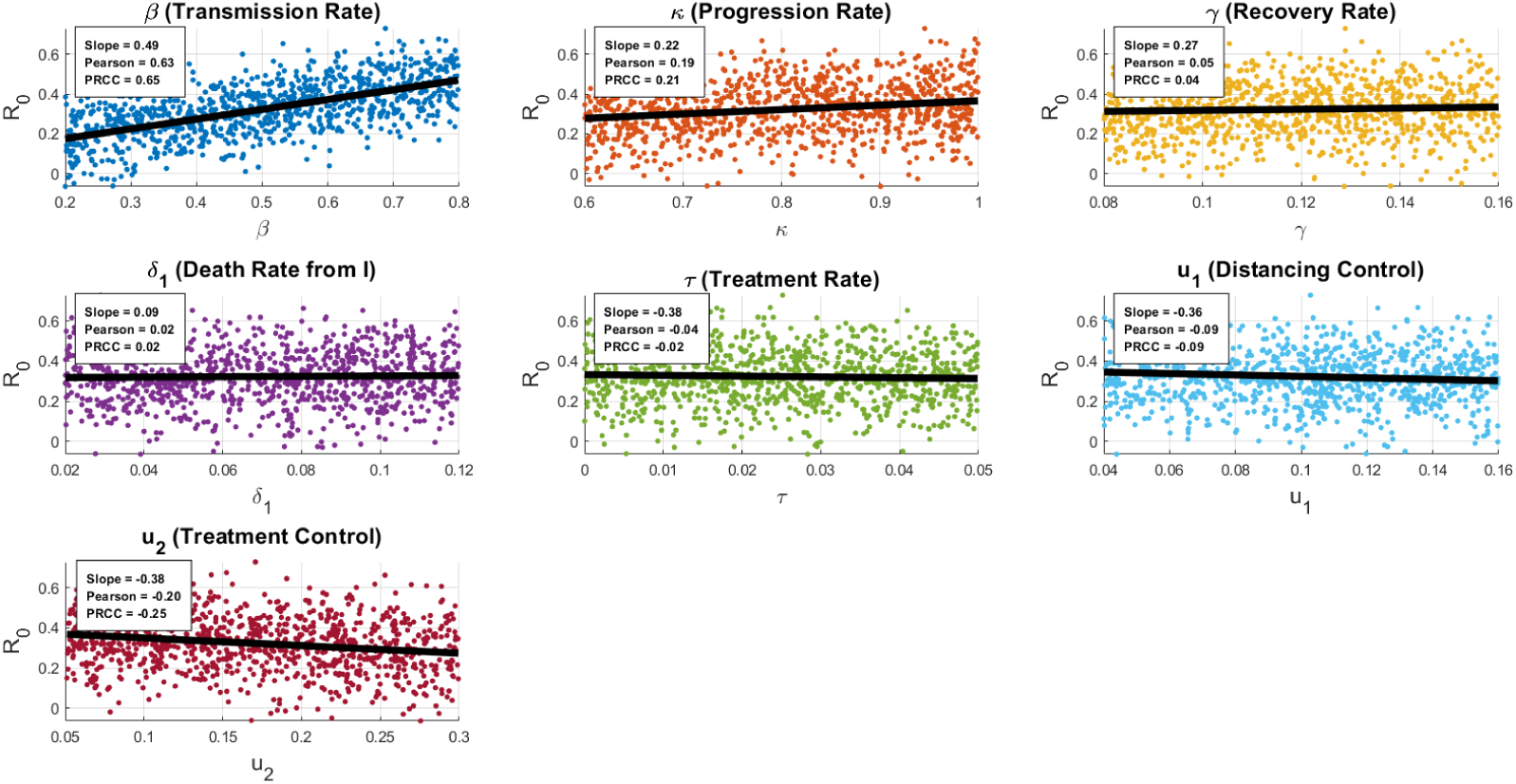
Regression, Pearson and PRCC analysis of model parameters by scatter diagram.

Other parameters such as the disease-induced death rate from the infected class (*δ*_1_) showed minimal influence on *R*_0_, with slope (0.09), Pearson (0.02), and PRCC (0.02) values close to zero. In summary, the most critical parameters for TB control based on this analysis are *β* and *u*_2_, which had the highest opposing sensitivities. The Table 3 below summarizes the key sensitivity indicators:

**Table 3:** Sensitivity Metrics for SEITR Model Parameters.

| Parameter | Slope | Pearson | PRCC | Interpretation |
| --- | --- | --- | --- | --- |
| $\beta$ | +0.49 | +0.63 | +0.65 | Strong positive influence on $R_0$ |
| $\kappa$ | +0.22 | +0.19 | +0.21 | Moderate positive influence |
| $\gamma$ | +0.27 | +0.05 | +0.04 | Weak, likely noise |
| $\delta_1$ | +0.09 | +0.02 | +0.02 | Negligible impact |
| $\tau$ | -0.38 | -0.04 | -0.02 | Weak inverse effect |
| $u_1$ | -0.36 | -0.09 | -0.09 | Mild negative effect |
| $u_2$ | -0.38 | -0.20 | -0.25 | Most effective control parameter |

Figure 5 displays the Partial Rank Correlation Coefficient (PRCC) values for different parameters affecting the five groups in a TB SEITR model that includes distancing and treatment controls. Each bar graph shows how sensitive each compartment is to changes in parameters like the transmission rate (*β*), progression rate (*κ*), recovery rate (*γ*), death rates (*µ*, *δ*_1_, *δ*_2_), control strategies (*u*_1_*, u*_2_), and recruitment rate (Π). Parameters with bars that are close to 1 or −1 have the most effect. If the value is positive, raising the parameter will raise the number in that group. If the value is negative, lowering the parameter will lower the number. From an epidemiological standpoint, the PRCC analysis elucidates the parameters that exert the most significant influence on the model’s compartments. A high positive PRCC value for the transmission rate *β* and the progression rate *κ* in the susceptible and exposed populations indicates that these parameters significantly influence the transition of individuals into infection. Distancing control *u*_1_, treatment control *u*_2_, and disease-related death rates show strong negative correlations for the infected class, indicating their efficacy in reducing active infection. Treatment effort *u*_2_ and recovery rate *γ* are found to have a significant impact on the treated and recovered compartments, respectively, underscoring the vital significance of maximizing contact reduction, treatment efficiency, and recovery in tuberculosis control.

**Figure 5:**
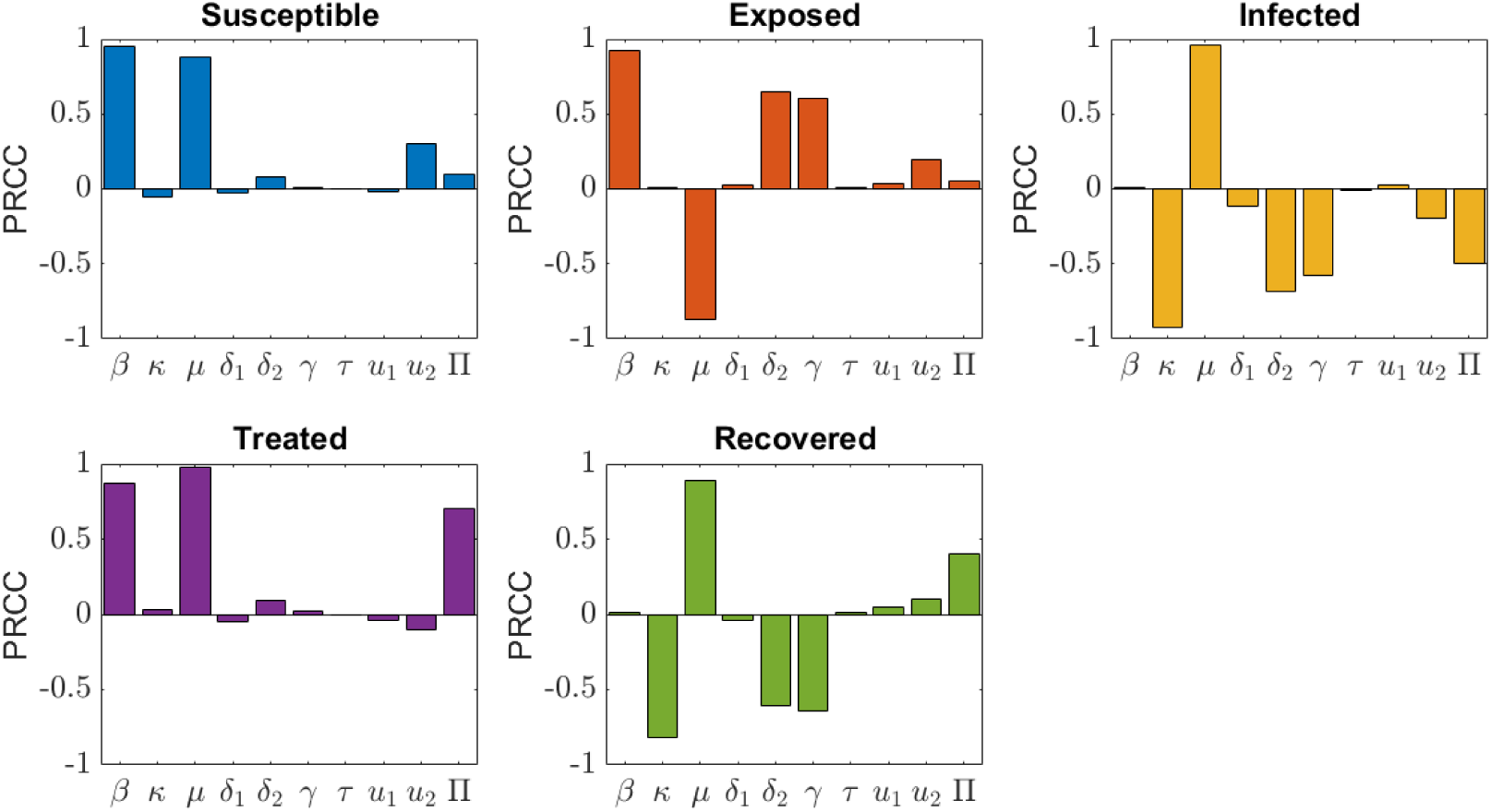
PRCC indices of all parameters for each compartments of the model.

Figures 6 and 7 illustrate how the basic reproduction number (*R*_0_) changes in response to variations in the treatment parameter *τ* and the transmission rate *β* within the SEITR model, where values greater than one suggest disease persistence and values less than one indicate elimination. An inverse and nonlinear relationship is evident between *R*_0_ and *τ* , with elevated reproduction levels present at low *τ* values, which quickly decline toward nearzero as *τ* increases, signifying improved epidemic management. A direct and roughly linear relationship is observed between *R*_0_ and *β*, where an increase in transmission intensity results in a proportional rise in the reproduction number. When *β* reaches a certain point, the epidemic threshold is crossed, and continuous transmission is expected. These findings underscore that both efficient transmission reduction and the improvement of treatment-related processes are essential for reducing *R*_0_ below the critical threshold and achieving disease control.

**Figure 6:**
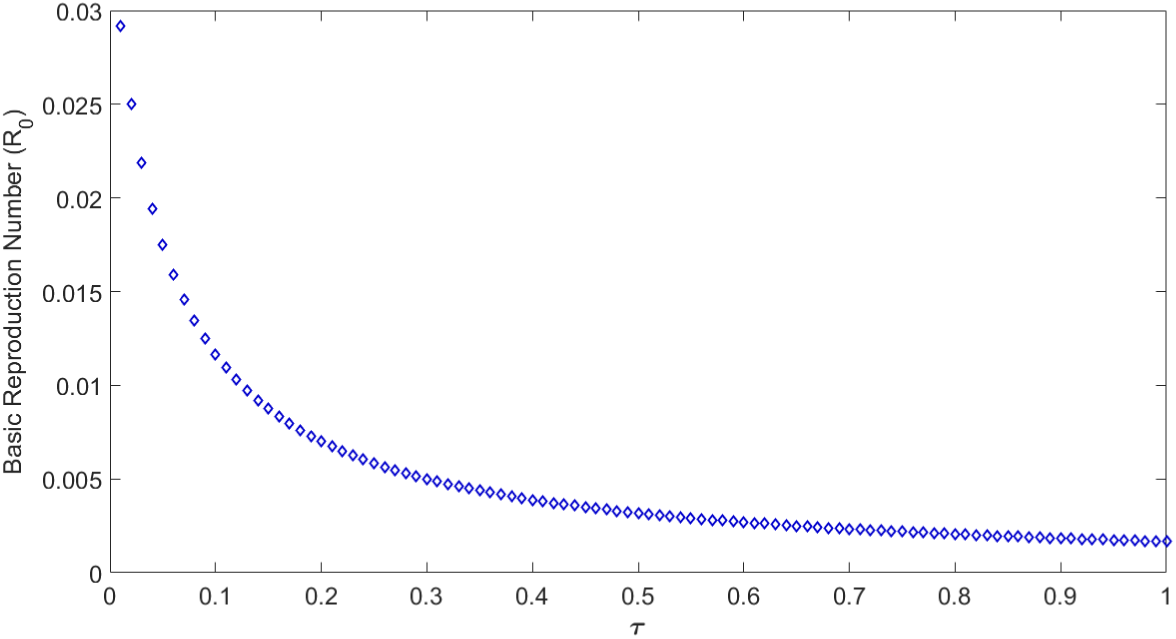
Plot of parameter *τ* with the basic reproduction number (*R*_0_).

**Figure 7:**
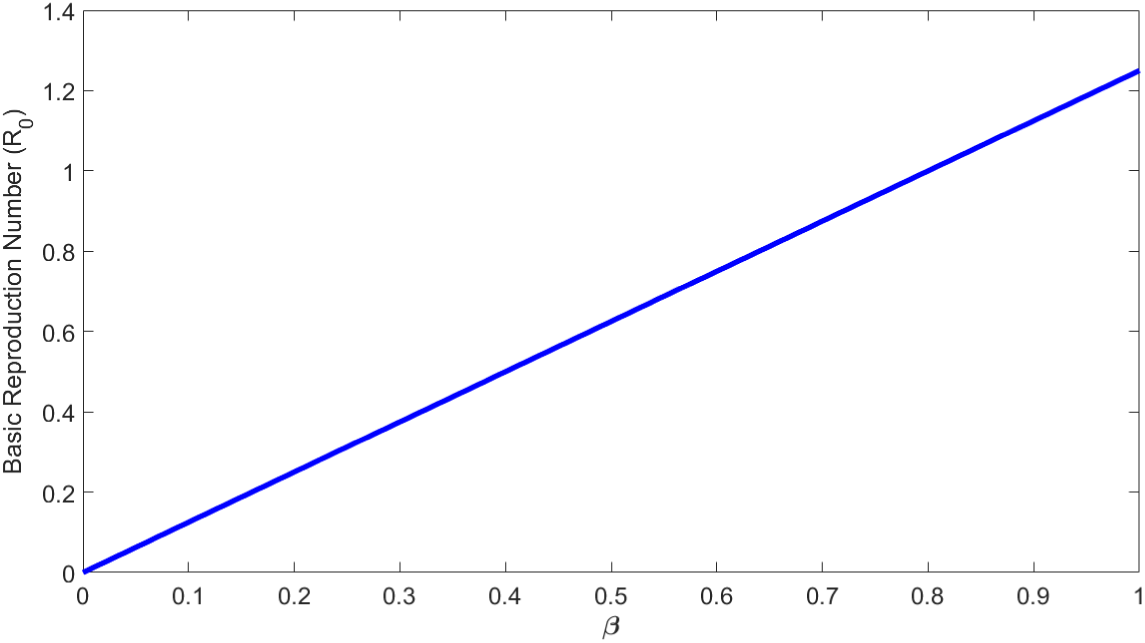
Plot of parameter *β* with the basic reproduction number (*R*_0_).

### 5.3 Control Panels Profile

The control panel profile is used to show how important intervention parameters change and work together in the proposed modeling structure. We look at how control measures affect system behavior and how they can help with disease control by showing how they work. The following graphs show the controls that were used in the model: Figures 8(a), 8(b), and 8(c) show the best time-dependent ways to control an epidemic. They show prevention (*u*_1_, labeled as Distancing) and treatment (*u*_2_, labeled as Treatment) efforts over a 200-day period. The graphs show that a good control strategy needs both high levels of distancing and treatment at first. For the first 60 days, distancing efforts (*u*_1_) should be kept at 0.4, then quickly drop. This suggests that there should be a strong push for prevention early on. In contrast, treatment efforts (*u*_2_) are maintained at a higher level of 0.7 for a longer duration, approximately 100 days, before gradually declining. This indicates a phased approach where early and strong prevention measures are crucial, followed by a sustained and then gradually reduced treatment intervention, to effectively manage the epidemic as modeled by an SEITR compartmental system. The weight parameters (*a*_1_*, a*_2_*, a*_3_*, a*_4_*, a*_5_) influence the relative importance of different objectives in the control problem. Such as, a higher weight on treatment might lead to a stronger emphasis on treatment interventions. By comparing the control strategies across the three figures, we can see how changes in the weight parameters affect the optimal control decisions.

**Figure 8:**
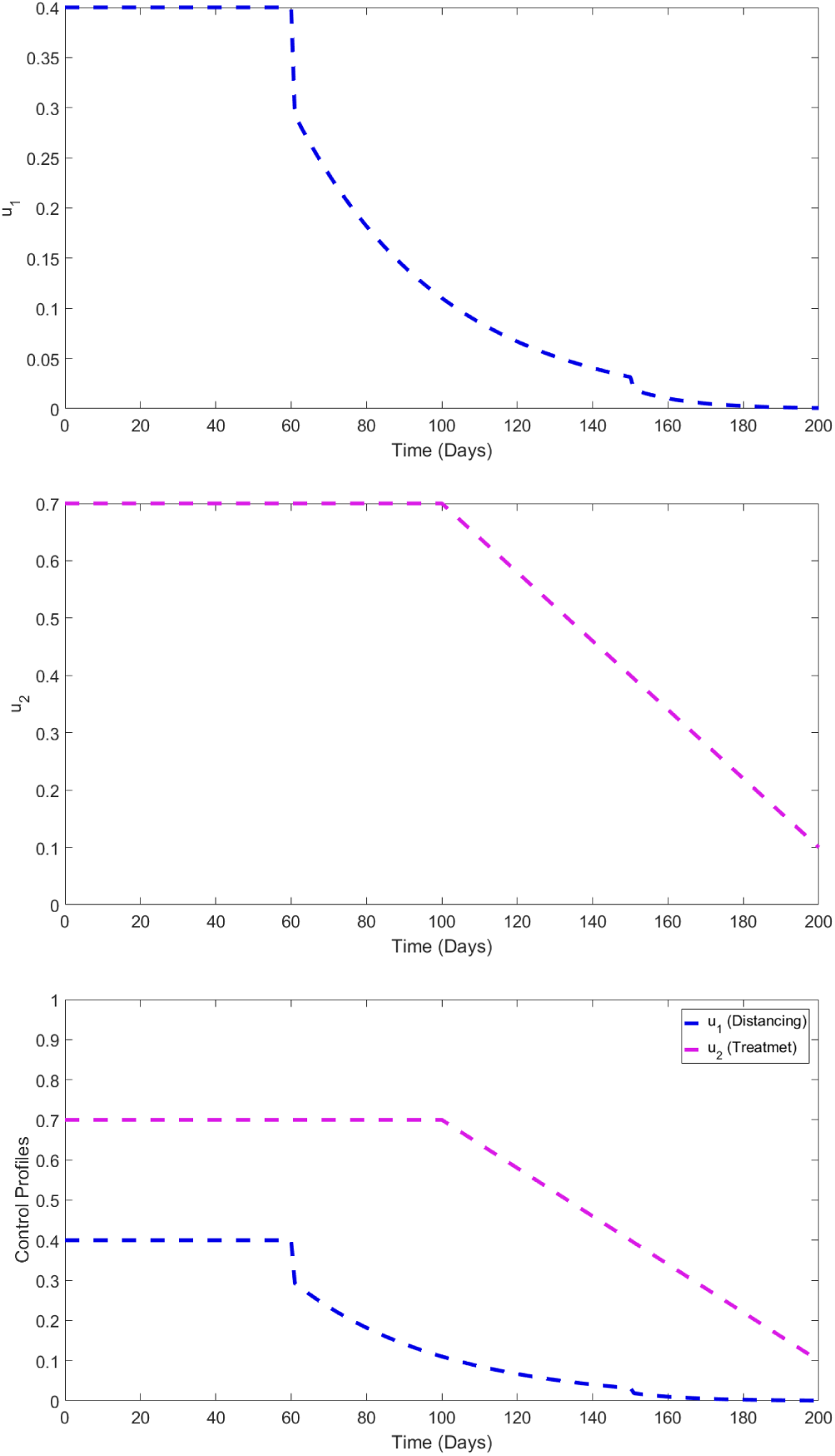
Different controls panels (a),(b),(c) using different weight parameters.

The Figure 9 shows the results of a tuberculosis (TB) mathematical model where different strategies for controlling the disease are applied over time. It compares four scenarios for controlling the spread of TB based on two types of controls: distancing control (*u*_1_) and treatment control (*u*_2_). In this figure, the Green dashed line (*u*_1_ = 0*, u*_2_ = 0) represents no controls are applied (that means no distancing or treatment). In this case, the number of infected individuals keeps rising steadily over time. This represents the worst-case scenario where the disease spreads unboundedly. The Red dashed line (*u*_1_ ≠ 0*, u*_2_ = 0) represents only distancing control is applied, but there is no treatment control. In this case, the number of infections decreases compared to the green line, but not as effectively as other strategies. Distancing helps to reduce the spread, but it cannot completely control the outbreak.

**Figure 9:**
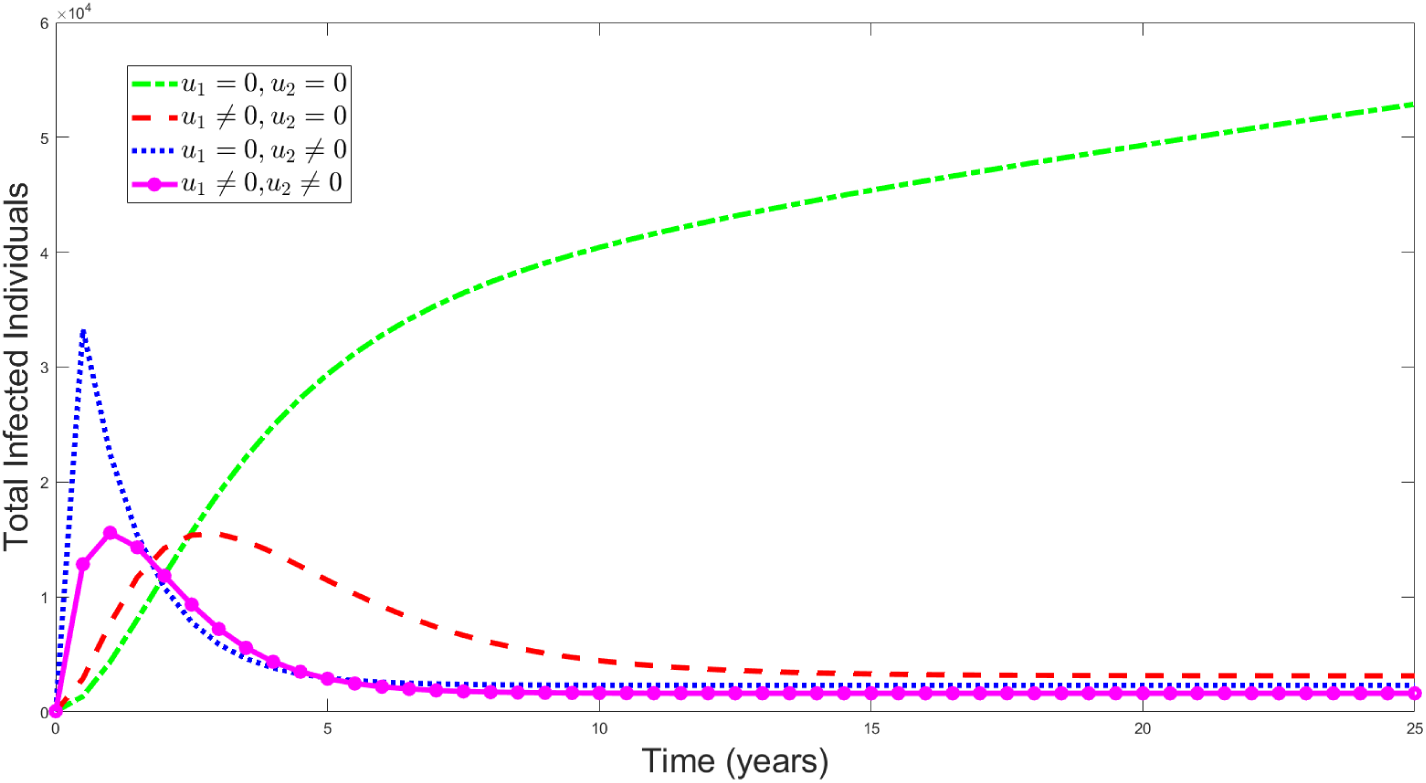
The density of total infected individuals analyzed with and without control measures.

The Blue dotted line (*u*_1_ = 0*, u*_2_ ≠ 0) represents only treatment control is applied, but no distancing control. In this case, the number of infections initially rises, but treatment eventually causes the infected population to decline significantly. This approach is more effective than distancing alone. Magenta line with circles (*u*_1_ ≠ 0*, u*_2_ ≠ 0) represents both distancing and treatment controls are applied together. In this case, this strategy yields the best results. Infections are reduced rapidly and sustainably, demonstrating that combining distancing and treatment is the most effective way to control TB.

### 5.4 Curve Fitting of Infected Cases with Model Prediction with Projection

In Figure 10, titled TB Case Projection with Fitted Curve (2010–2030) visually represents the alignment between real TB case data and model-based predictions using a SEITR (Susceptible–Exposed–Infectious–Treated–Recovered) epidemiological framework. In this figure, the green bars correspond to the observed TB case data from 2010 to 2025 [**?**], based on surveillance and reporting. The red bars project TB cases from 2026 to 2030 using the SEITR model, which incorporates disease progression and control strategies such as treatment and social distancing. A smooth blue curve has been fitted across the entire dataset using nonlinear regression or similar techniques to capture the trend of infection spread over time. The curve fitting serves as a validation tool to assess how accurately the SEITR model captures the real behavior of TB transmission dynamics. The fitted curve shows that the number of TB cases steadily rises until it reaches a peak around 2025, after which it slowly falls during the forecast period. This pattern shows how actions modeled in the system, such higher treatment rates and behavioral adjustments like distance, have an effect. The fact that the curve and the real data (green bars) are so similar during the years studied shows that the model is very stable, which means it will continue to be useful for tracking population-level TB changes over time.

**Figure 10:**
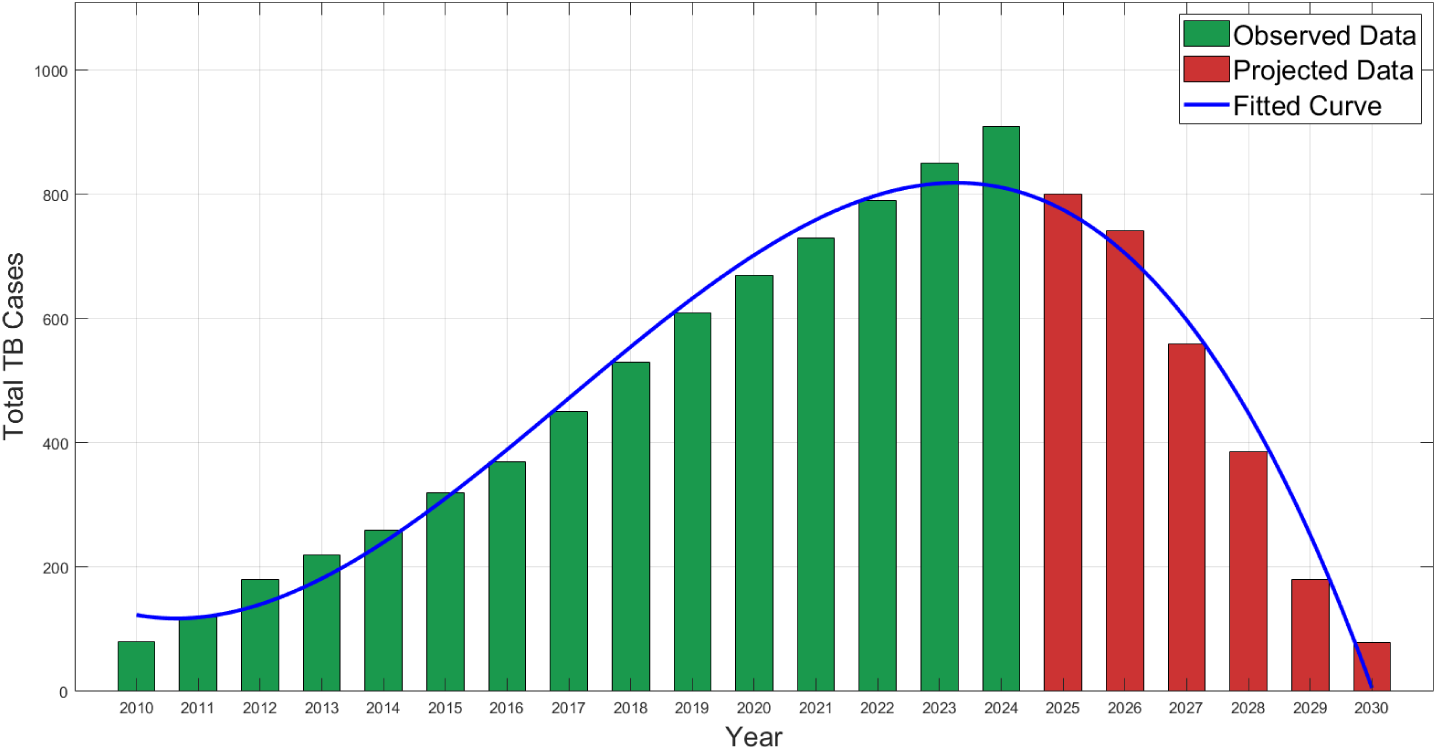
Curve fitting of infected cases with model prediction with projection up to 2030.

The model is also quite sensitive to control factors, as seen by the red bars showing a decrease in anticipated cases. This means that good treatment and distance can greatly lower the number of TB cases by 2030. This illustrates that mathematical modeling not only enhances comprehension of illness patterns but also functions as a forecasting instrument for policymakers.

Figure 11 shows how model-generated trends from 2010 to 2025 compare to reported occurrences of TB. The red plus signs depict real TB case data from 2010 to 2025, which shows a steady growth from fewer than 100 cases to almost 1000. The orange line, which shows the model without any controls, follows this pattern quite closely and keeps going up fast. This means that the model is correct when it comes to how TB spreads without any controls. The green line, which shows the model with controls (such treatment and distancing), goes up considerably more slowly and cuts down on the number of TB cases. The space between the green and orange lines clearly stops at about 700 cases in 2025. This slower rise illustrates that the interventions are working and how big of a difference control methods can make. The figure shows that timely controls can significantly lower the number of TB cases over time.

**Figure 11:**
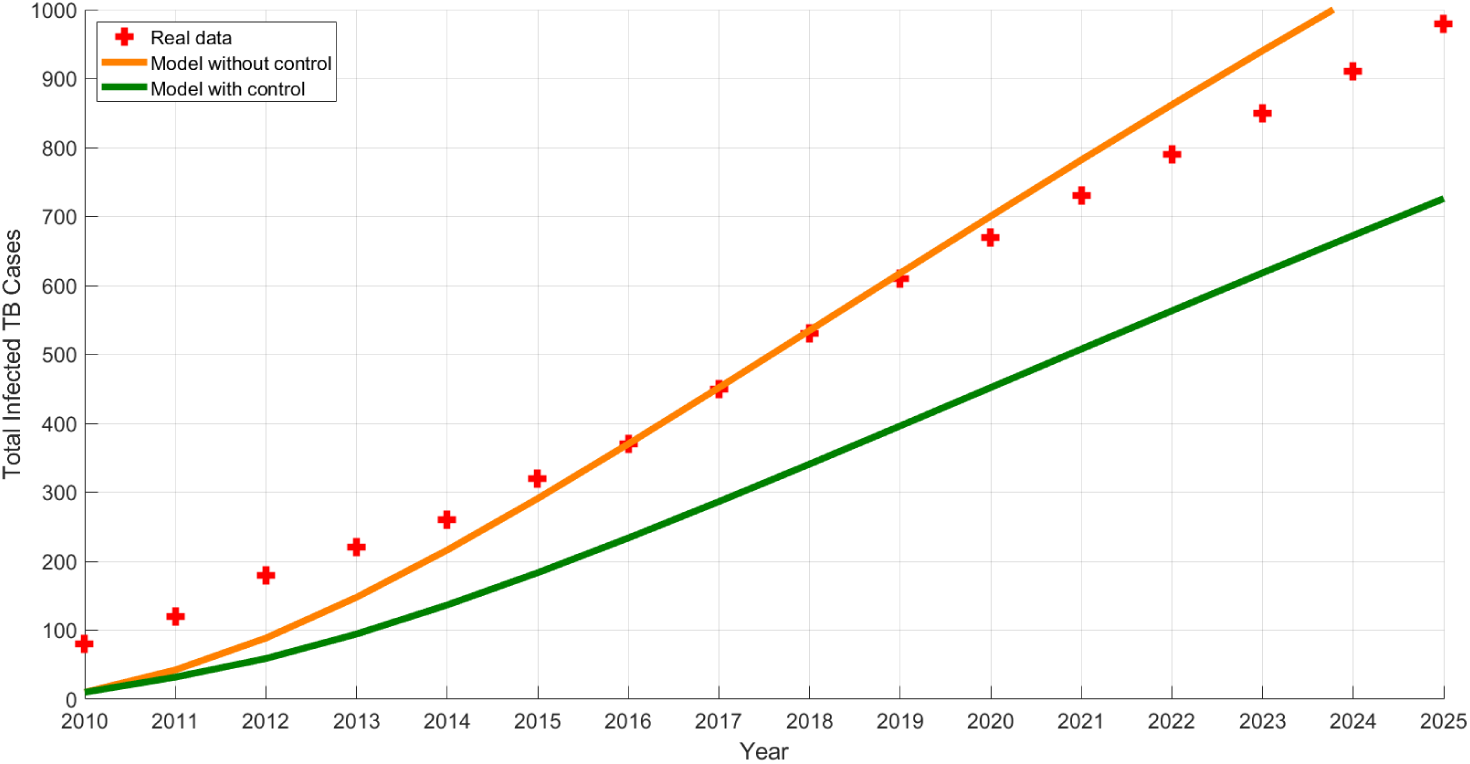
Model Fit to Real Data (2010-2025) of total TB cases assessed with and without control measures.

## 6 Novelty of the Work

A composite analytical methodology is proposed that integrates control-oriented stability decomposition with normalized forward sensitivity indexing to analyze dual intervention effects in a tuberculosis model. This method produces novel analytical threshold relations and dominance criteria that differentiate the independent, competitive, and cooperative effects of the two controls on system equilibria. The outcomes yield novel criteria for identifying optimal intervention strategies that reduce disease persistence while ensuring dynamical stability.

## 7 Conclusion

In this research, an SEITR mathematical model has been developed to examine the transmission of tuberculosis and evaluate the efficacy of distancing and treatment interventions. It is well known that transmission and progression parameters have a big effect on the basic reproduction number. This highlights the main factors that keep a disease going. The model’s stability is confirmed through analysis, which guarantees that the suggested structure accurately represents long-term population dynamics. To lower infection rates, optimal control theory is used. The following are the epidemiological and control-related effects of the main findings:

- Evidence demonstrates that diminishing the effective contact rate (*β*) through distancing measures substantially inhibits the transmission of new infections.
- Better treatment programs have been shown to speed up recovery and lower the death rate among people who are infected.
- The simultaneous implementation of distancing (*u*_1_) and treatment (*u*_2_) strategies has proven to be more effective than methods relying exclusively on a single intervention.
- It has been recognized that timely and ongoing implementation of control measures is crucial for maintaining disease stability.
- The model’s alignment with actual data has been validated, reinforcing its importance for public health strategy development.

In conclusion, it is evident that combining behavioral and medical interventions offers the most effective approach for controlling tuberculosis. The significance of ongoing public awareness, access to health care, and adherence to treatment are emphasized by the outcomes of the models. Mathematical modeling has proven to be an effective decision-support tool for enhancing intervention design and resource distribution. These results bolster evidence-based policy making focused on the long-term reduction of diseases.

## CRediT Author Statement

**M. A. Salek**: Data curation, Investigation, Methodology, Software, Validation, Writing-original draft; **J. Nayeem**: Conceptualization, Formal analysis, Methodology, Funding, writing original draft; **Muhammad Sajjad Hossain**: Conceptualization, Administration, Methodology, Resources, Supervision, writing review and editing; **M. H. Kabir**: Visualization, Supervision, Methodology, writing review and editing. All authors have read and agreed to the published version of the manuscript.

## Funding

This research received the University Grand Commission (UGC) of Bangladesh funding.

## Data Availability

www.ntp.gov.bd/wp-content/uploads/2024/01/Annual-Report-2022.pdf

## Acknowledgments

All authors want to express thankfulness to everyone for contribution to accomplish this research work.

## Conflicts Of Interest

The authors declare no conflict of interest.

## Appendix

Reference values and the biological meaning of the SEITR model are given in Table 1.

## Notes

### Competing Interest Statement

The authors have declared no competing interest.

### Funding Statement

Name of granting agency: University Grand Commission Bangladesh. Grant number: PhD Fellowship 2022-2023, serial no. 52. Description of the funder’s role: Provide full fund for any publication

